# Association between age at menopause and 14-year cognitive trajectories in the English Longitudinal Study of Ageing

**DOI:** 10.64898/2026.01.27.26344915

**Authors:** Qingmei Chen, Jianye Dong, Haibin Li, Ying Chen, Guo-Chong Chen, Jianian Hua

## Abstract

**Introduction:** The association between age at menopause and long-term cognitive decline remains largely unknown. We examined the association between age at menopause and 14-year cognitive trajectories among UK population.

**Methods:** This community-based study enrolled 4,082 postmenopausal women (mean age, 64.2±10.7 years) with baseline cognition and at least one follow-up assessment from the English Longitudinal Study of Ageing (ELSA) Waves 3–10. Age at menopause was categorized as <40 (premature), 40–49, and ≥50 years (late), with ≥50 years as the reference. Cognitive function was measured by global cognition, which was a composite of three domain-specific tests: semantic fluency, memory, and orientation. Multivariate-adjusted linear mixed models were employed to estimate the longitudinal associations.

**Results:** Compared with late menopause, menopause at 40–49 years was not associated with lower baseline global cognition or faster long-term decline in global cognition. Premature was associated with lower baseline global cognition (β=-0.218 SD, 95% CI -0.313 to -0.123, *p*<0.001), with consistent finding across semantic fluency, memory, and orientation tests. However, premature menopause was not significantly related to a faster rate of decline in global cognition (β=-0.010 SD/year, 95% CI -0.022 to 0.002, *p*=0.112). In domain-specific tests, premature menopause was associated with faster decline in memory test (β=-0.011 SD/year, 95% CI -0.021 to -0.001, *p*=0.027), while menopause at 40–49 years was associated with faster decline in semantic fluency test (β=-0.007 SD/year, 95% CI -0.013 to -0.001, *p*=0.015). Surgical menopause and hormone replacement therapy did not modify the association between premature menopause and global cognition.

**Conclusion:** Premature menopause was associated with poorer cognitive performance later in life, whereas long-term global cognitive decline was broadly similar across menopause-age groups. These findings suggest that the cognitive impact of premature menopause may be concentrated in earlier postmenopausal years, highlighting the importance of monitoring cognitive change and implementing preventive strategies soon after menopause.

## 1. Introduction

Women have twice the prevalence of Alzheimer’s’ Dementia (AD) compared to men. [1] Growing evidence suggests that this sex disparity may partly attribute to the decline in endogenous estrogen production following menopause, due to the neuroprotective effects of estrogen.[2, 3] Age at menopause reflects the cumulative duration of exposure to estrogen. Accordingly, earlier menopause may indicate a shorter period of estrogen-related neuroprotection and represents a potentially important target for strategies to preserve cognitive function and reduce the burden of dementia.

However, prior studies have reported inconsistent associations between earlier age at menopause and risk of dementia. While some studies observed a higher dementia risk among women with earlier menopause, others found no significant relationship. [4-6] These discrepancies underscore the need to clarify the relationship between early menopause and later-life cognitive health.

Long-term cognitive decline may serve as a more informative endpoint than dementia, as it captures gradual changes in cognitive function which can impair daily activities and quality of life, even before the onset of mild cognitive impairment (MCI) or dementia.[7] Importantly, the early symptomatic stage may offer a critical window for intervention, including management of modifiable risk factors, symptomatic therapies (e.g., donepezil or memantine) for cognitive impairment, and disease-modifying treatments (e.g., aducanumab or lecanemab) for early AD. Therefore, understanding the association between age at menopause and cognitive decline may provide actionable insights for protecting women’s cognitive health.

Despite sustained interest, the impact of age at menopause on the rate of cognitive decline remains unclear. Existing studies focused on the short-term cognitive changes or cross-sectional cognitive performance at one single time point, which are more vulnerable to reverse causation and cannot capture within-person trajectories.[8, 9] In contrast, long-term trajectories of cognitive decline provide a more robust framework to evaluate the relationship. Moreover, hormone replacement therapy (HRT) use and surgically induced menopause are more common among women with earlier menopause, yet their roles in cognitive decline and their potential to modify the associations between menopause and cognition remain uncertain.

Using data from the English Longitudinal Study of Ageing (ELSA), we aimed to explore the association between age at menopause and long-term cognitive decline and examine the mediating effect of menopause-related factors, such as HRT use.

## 2. Methods

### 2.1. Study participants

We extracted data from the ELSA cohort. Details of this cohort are described elsewhere.[10] Briefly, the ELSA is a population-based observational study. The Wave 1 survey of ELSA was performed between 2002 and 2003, recruiting 17,708 community-dwelling participants, and the ongoing follow-up survey collected every 2 years. Ethical approval for ELSA was approved by the London Multicenter Research Ethics Committee (MREC/01/2/91). Each participant provided written informed consent.

For this study, we included the Wave 3 to Wave 10 survey because the menopause data was available since Wave 3 (n=9,771). We excluded 309 participants missing cognitive data at baseline, 65 reporting previously diagnosed dementia, 539 women without data on age at menopause, 126 women still menstruating, 71 women with unlikely age of menopause (6 [0.1%] <20 years and 65 [1.4%] >60 years), and 252 participants missing baseline covariates.[11] Among the 8,409 participants with complete data at baseline, we further exclude 988 participants who did not receive at least one cognitive test during the follow-up. The flow chart pf sample selection is shown in Figure 1.

**Figure 1.**
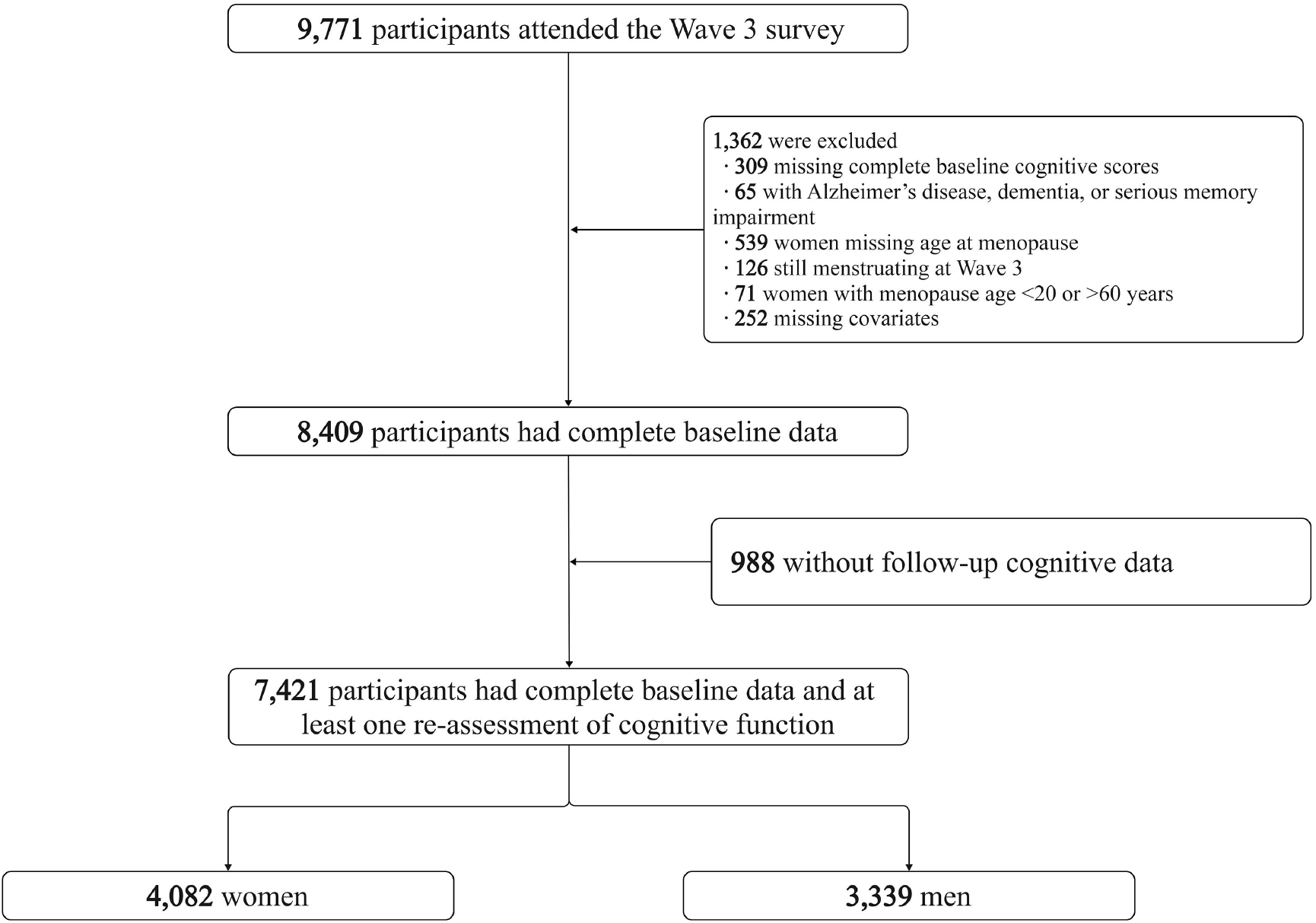
Flowchart of sample selection.

### 2.2. Age at menopause and reproductive factors

Women were asked whether they had menstruated in the last 12 months, the year of their last menstrual period, and the age at menopause. We classified age at menopause into <40 (premature menopause), 40–49, and ≥50 (late menopause) years according the Women’s Health Initiative Observational Study.[12] We used women ≥50 years as the reference group, as they represent the longest exposure to the protective hormone. [4]

We performed subgroup analysis for natural/surgical menopause and HRT use (yes/ no). Years of menstruating was calculated as the intervals between age at menarche and age at menopause.

### 2.3. Cognitive function

We generate a global cognitive z score as the composite of three tests: semantic fluency, memory, and orientation. Semantic fluency test asked participants to name as many animals as possible in one minute (score range: 0–50), reflecting the executive function. In memory test, the interviewer reads 10 unrelated words, asking participants to recall them immediately and after a few minutes (the sum score of immediate recall and delayed recall ranges from 0–20). The orientation test asked participants to name the year, month, data, and day of the week (score range: 0–4). We firstly create *z*-scores by the mean and SD at baseline for three tests separately. A z score of 1.000 means 1 SD higher than the mean score at baseline. Secondly, we create a global cognitive z score by the mean of three scores and then create the z score in the same manner.

### 2.4. Covariates

We included baseline covariates which were associated with cognitive function or age at menopause, including age (continuous variable), age squared, body mass index (kg/ m^2^), education level (less than o-level, o-level or equivalent, or higher than a-level), smoking status (current, ever, or never), alcohol drinking (at least once per week or not), physical activity(inactive, mild, moderate, and higher), depression (yes or no), hypertension, diabetes, stroke, and cancer. Details of variables are shown in Table S1.

### 2.5. Statistical analysis

Baseline variables were described as mean (SD) for continuous variables and No. (%) for categorical variables, by age at menopause and sex. We compared the difference through *t* test for continuous variables and χ^2^ test for categorical variables, with late menopause as the reference group.

We constructed linear mixed models to compare the cognitive function between different age at menopause. The models could take into account repeated measurements of cognitive function and missing outcomes data.[13, 14] The model included fixed effects and random effects. Fixed effects include intercept, time (reflecting the cognitive decline rate of the reference group), age at menopause (reflecting the difference in baseline cognitive scores across each group), age at menopause × time (reflecting the difference in cognitive decline rate across each group), age × time, and covariates. Random effects included intercept and time, to account for variability in baseline cognitive scores and rate of cognitive decline. The visualization results are exhibited in Figure 2 and Figure S1.[15] Furthermore, we applied restricted cubic spline (RCS) to test the dose–response relationship between age at menopause, reproductive period, and time to baseline with cognitive function. The rate of cognitive decline in RCS was calculated by subtracting the cognitive score at the last visit from score at the first visit and dividing it by the follow-up duration (years) for each participant.[16]

**Figure 2.**
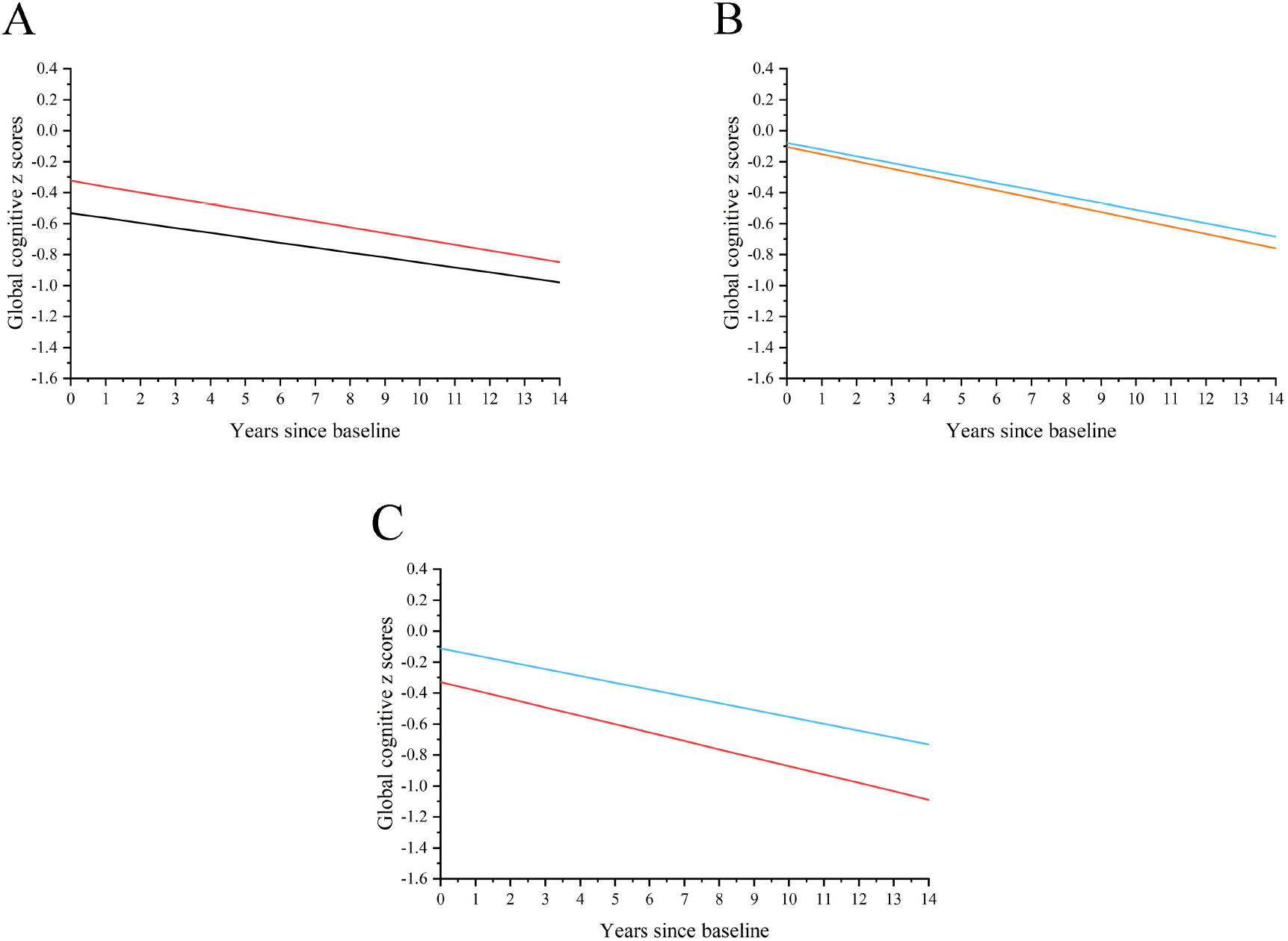
Trajectories of cognitive function by sex and age at menopause. The values were based on the linear mixed models. After adjustment for baseline age, age squared, age×time, BMI, marital status, education level, smoking, drinking, physical activity, depression, hypertension, diabetes, stroke, and cancer. Baseline covariates were set to the most common characteristics: age 65 years, BMI 28, married, education less than o-level, low physical activity, current smoking, alcohol drinking at least once per week, with hypertension, without depression, diabetes, stroke or cancer. Random effects were set to 0. (A) Black line: men; red line: women. (B) Blue line: women with age at menopause ≥50 years; orange line: women with age at menopause between 40–49 years. (C) Blue line: women with age at menopause ≥50 years; red line: women with age at menopause <40 years.

We conducted a set of subgroup analysis to explore whether our main results varied across subgroups. All subgroups were reported be associated with age at menopause or cognitive decline, including time from menopause to baseline, natural menopause/ surgery, use of hormone replacement therapy, baseline age, educational level, marital status, body mass index, and smoking status. The main analysis was conducted in each subgroup separately, and an integrated model in which the fixed effect included intercept, time, age at menopause, age at menopause ×time, subgroup, subgroup×time, subgroup× age at menopause, subgroup× age at menopause ×time, and covariates. The *p* value for the interaction term “subgroup ×age at menopause× time” reflected whether the subgroup modified the effect of age at menopause on cognitive decline.

We conducted a serious of sensitivity analysis by repeating our primary analysis: a) only include participants who completed cognitive tests in at least 6 waves, which tended to show the trends in relatively healthier participants; b) classify age at menopause into tertiles; (c) include participants who were ≥55 years old at baseline to avoid immortal time bias; (d) exclude participants who menopaused within two years before baseline, considering the high incidence of depression and estrogen deprivation perimenopause; (e) exclude those with stroke history at baseline or developed stroke during the follow-up, considering the strong association between stroke and cognitive decline.

All analyses were conducted using SAS version 9.4M7 and R (v4.4). A two-sided *p* value of <0.05 was considered to indicate statistical significance.

## 3. Results

We ultimately included 4,082 females and 3,339 males in the study. Among females, 2,357 (58.2%) experienced late menopause, 1,420 (34.8%) experienced menopause between 40–50 years, and 305 (7.5%) experienced had premature menopause. The mean (SD) age at menopause was 48.9 (6.0) years. Each menopause-age subgroup received a mean of 4.9–5.1 cognitive assessments (Table S2). The distribution of age at menopause is shown in Figure 3. Compared with females experiencing late menopause, those with menopause <40 years and between 40–49 years were younger at baseline, were more likely to be current smokers, were less likely to drink alcohol, and had higher proportion of depression and hypertension. In addition, compared with late menopause, premature menopause was associated with higher BMI, less physical activity, less cognitive scores in each test, and a higher prevalence of stroke.

**Figure 3.**
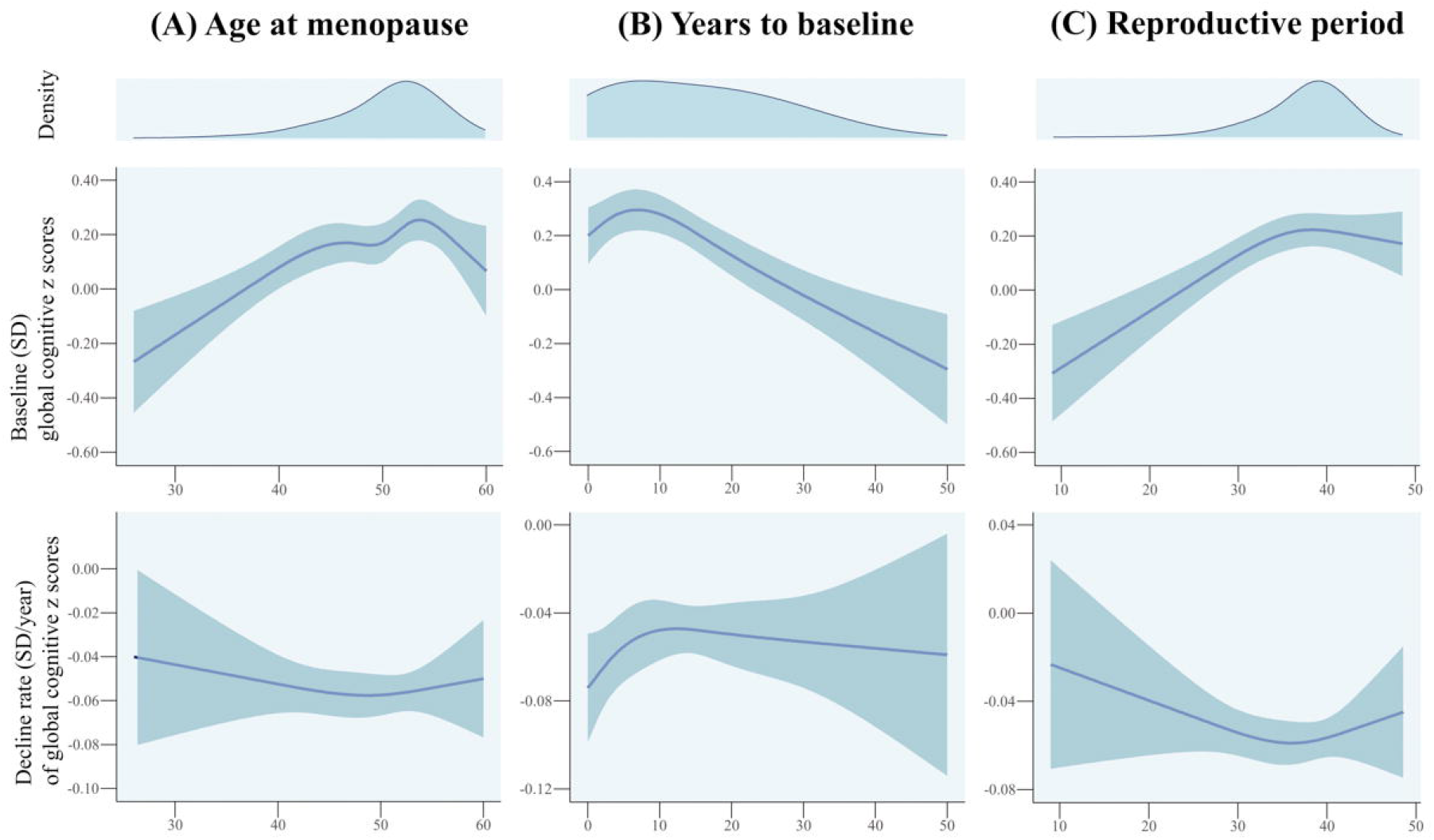
Dose–response associations between age at menopause, years from menopause to baseline, and reproductive period with baseline cognitive function and rate of cognitive decline during follow-up. Shaded areas represent 95% CI. Baseline cognitive function was adjusted for baseline age, age squared, age×time, BMI, marital status, education level, smoking, drinking, physical activity, depression, hypertension, diabetes, stroke, and cancer. Cognitive decline was adjusted for age.

Compared with females with late menopause, males were younger, more likely to be married, higher educated, smoking, drinking, and engaging in physical activity, and a higher prevalence of hypertension and diabetes, lower global cognitive z score, and a lower prevalence of depression (Table 1).

**Table 1.** Baseline characteristics by age at menopause and sex.

|  | Women |  |  | Men |
| --- | --- | --- | --- | --- |
|  | <40 years<br>(n=305, 7.5%) | 40–49 years<br>(n=1420,<br>34.8%) | ≥50 years<br>(n=2357,<br>58.2%) | (n=3339) |
| Age, mean (SD), years | 61.8 (11.2)* | 63.2 (11.2)* | 65.1 (10.2) | 64.2 (10.0)* |
| Body mass index, mean (SD), kg/m<sup>2</sup> | 28.8 (5.5)* | 28.1 (5.2) | 27.9 (5.1) | 28.0 (4.2) |
| Married or cohabitation | 201 (65.9) | 921 (64.9) | 1524 (64.7) | 2661 (79.7)* |
| Education level |  |  |  |  |
| Less than o-level | 138 (45.2) | 646 (45.5) | 1009 (42.8) | 1120 (33.5)* |
| O-level or equivalent | 96 (31.5) | 422 (29.7) | 701 (29.7) | 804 (24.1)* |
| Higher than a-level | 71 (23.3) | 352 (24.8) | 647 (27.5) | 1415 (42.4)* |
| Smoking status |  |  |  |  |
| Current smoking | 82 (26.9)* | 247 (17.4)* | 299 (12.7) | 490 (14.7)* |
| Ex smoker | 116 (38.0)* | 550 (38.7)* | 948 (40.2) | 1817 (54.4)* |
| Never | 107 (35.1)* | 623 (43.9)* | 1110 (47.1) | 1032 (30.9)* |
| Alcohol drinking at least once per week | 127 (41.6)* | 601 (42.3)* | 1142 (48.5) | 2105 (63.0)* |
| Physical activity |  |  |  |  |
| Inactive | 28 (9.2)* | 101 (7.1) | 132 (5.6) | 197 (5.9)* |
| Low | 52 (17.0)* | 234 (16.5) | 338 (14.3) | 297 (8.9)* |
| Moderate | 149 (48.9)* | 702 (49.4) | 1215 (51.5) | 1661 (49.7)* |
| High | 76 (24.9)* | 383 (27.0) | 672 (28.5) | 1184 (35.5)* |
| Depression | 83 (27.2)* | 256 (18.0)* | 352 (14.9) | 341 (10.2)* |
| Hypertension | 172 (56.4)* | 753 (53.0)# | 1153 (48.9) | 1751 (52.4)* |
| Diabetes | 19 (6.2) | 101 (7.1) | 169 (7.2) | 357 (10.7)* |
| Stroke | 20 (6.6)* | 56 (3.9) | 78 (3.3) | 128 (3.8) |
| Cancer | 13 (4.3) | 62 (4.4) | 100 (4.2) | 148 (4.4) |
| Memory score, mean (SD) | 10.5 (3.5)* | 10.9 (3.4) | 11.0 (3.5) | 10.2 (3.4)* |
| Executive score, mean (SD) | 19.2 (6.0)* | 20.6 (6.5) | 20.6 (6.5) | 20.9 (6.7)* |
| Orientation score, mean (SD) | 3.7 (0.7)* | 3.8 (0.5) | 3.8 (0.4) | 3.8 (0.5)* |
| Global cognitive z score, mean (SD) | -0.2 (1.1)* | 0.1 (1.0) | 0.1 (1.0) | -0.1 (1.0)* |

\* $p < 0.05$ , compared with the late menopause group.

When using linear mixed models to compare the cognitive trajectories between males and females, females had higher baseline global cognitive z scores. During follow-up, males showed a steeper cognitive decline (β=-0.006 SD/year, 95% CI -0.011 to -0.001, *p*<0.001), but their cognitive scores remained lower than those of females (Figure 2). Compared with females with late menopause, those with menopause between 40–50 years did not show significantly difference in baseline global cognitive scores or rate of global cognitive decline. For three domain-specific tests, menopause between 40–50 years was associated with faster long-term decline in semantic fluency test (β=-0.007 SD/year, 95% CI -0.013 to -0.001, *p*=0.015; Tables S3–S5 and Figure S1).

Compared with late menopause, premature menopause was significantly associated with lower baseline global cognitive z scores (β=-0.218 SD, 95% CI -0.313 to -0.123, *p*<0.001; Table 2). This result was consistent in all three tests of semantic fluency (β=-0.181 SD, 95% CI -0.276 to -0.085, *p*=0.005), memory (β=-0.133 SD, 95% CI -0.227 to -0.040, *p*=0.005), and orientation (β=-0.138 SD, 95% CI -0.236 to -0.039, *p* 0.006; Tables S3–S5 and Figure S1). However, premature menopause was not significantly related to a faster rate of decline in global cognition (β=-0.010 SD/year, 95% CI -0.022 to 0.002, *p*=0.112; Table 2). In the memory test, females with premature menopause exhibited steeper long-term cognitive decline (β=-0.011 SD/ year, 95% CI -0.021 to -0.001, *p*=0.027; Table S4).

**Table 2.** Difference in trajectories of cognitive function by sex and age at menopause^a^.

|  | Difference in baseline global cognitive z scores |  | Difference in decline rates of global cognitive z scores |  |
| --- | --- | --- | --- | --- |
|  | Coefficient (95% CI) | *p* value | Coefficient (95% CI) | *p* value |
| Sex |  |  |  |  |
| Male | Reference | Reference | Reference | Reference |
| Female | 0.208 (0.170, 0.246) | <0.001 | -0.006 (-0.011, -0.001) | 0.015 |
| Age at menopause |  |  |  |  |
| ≥50 y | Reference | Reference | Reference | Reference |
| 40–49 y | -0.026 (-0.077, 0.025) | 0.314 | -0.004 (-0.010, 0.003) | 0.311 |
| <40 y | -0.218 (-0.313, -0.123) | <0.001 | -0.010 (-0.022, 0.002) | 0.112 |

<sup>a</sup>Adjusted for baseline age, age squared, age×time, BMI, marital status, education level, smoking, drinking, physical activity, depression, hypertension, diabetes, stroke, and cancer.

The RCS analysis represented an inverted U-shaped association between age at menopause and baseline global cognitive z scores, in which, before approximately 50 to 60 years, earlier menopause was associated with lower baseline cognitive scores. However, no linear association was observed between age at menopause and the rate of long-term cognitive decline in RCS (Figure 3).

Across subgroups defined by years from menopause to baseline, menopause type (natural/surgery), and HRT use, premature menopause was associated with lower baseline cognitive function in most subgroups (Figure 4). Among females with natural menopause, menopause between 40–49 years (vs. late menopause) was associated with lower baseline cognitive function. Among those with 19–36 years from menopause to baseline, premature menopause was associated with faster long-term cognitive decline (β=-0.041 SD/year, 95% CI -0.081 to -0.001, *p*=0.047). A longer interval from menopause to baseline attenuated the negative effect of premature menopause on cognitive decline (*p* for interaction=0.040). RCS also indicated that longer time from menopause to baseline was associated with lower baseline cognitive function, but not with the rate of long-term decline (Figure 3). An inverted U-shaped association suggested that short reproductive period was associated with lower baseline cognitive function, but not with long-term cognitive decline.

**Figure 4.**
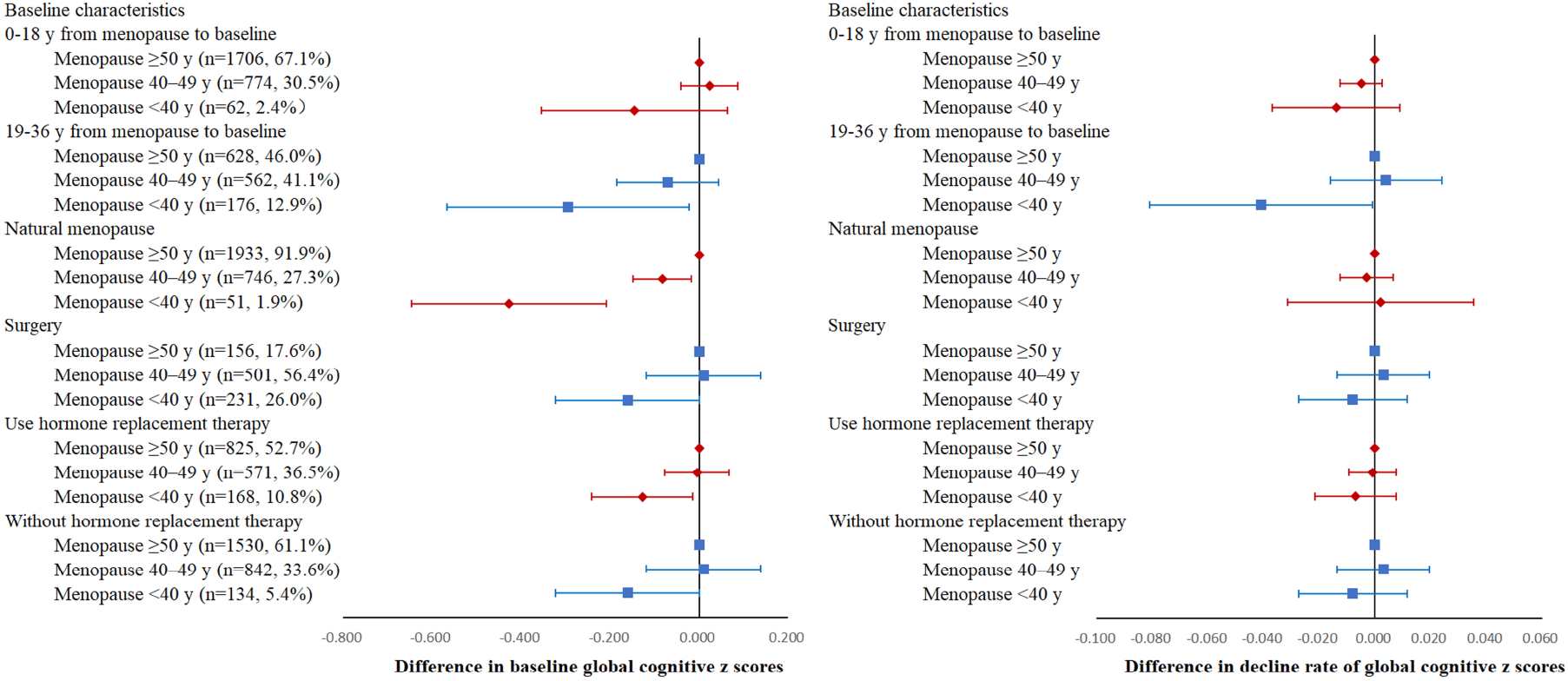
Associations between age at menopause and trajectories of cognitive function in subgroups according to years from menopause to baseline, HRT use, and surgical menopause. After adjustment for baseline age, age squared, age×time, BMI, marital status, education level, smoking, drinking, physical activity, depression, hypertension, diabetes, stroke, and cancer.

In subgroup analysis according to baseline age, education level, marital status, BMI, and smoking status, premature menopause was associated with lower cognitive function but was not significantly associated with faster long-term cognitive decline (Figure 5). The interaction term suggested that higher BMI attenuated the negative effect premature menopause on baseline cognitive score (*p*=0.022).

**Figure 5.**
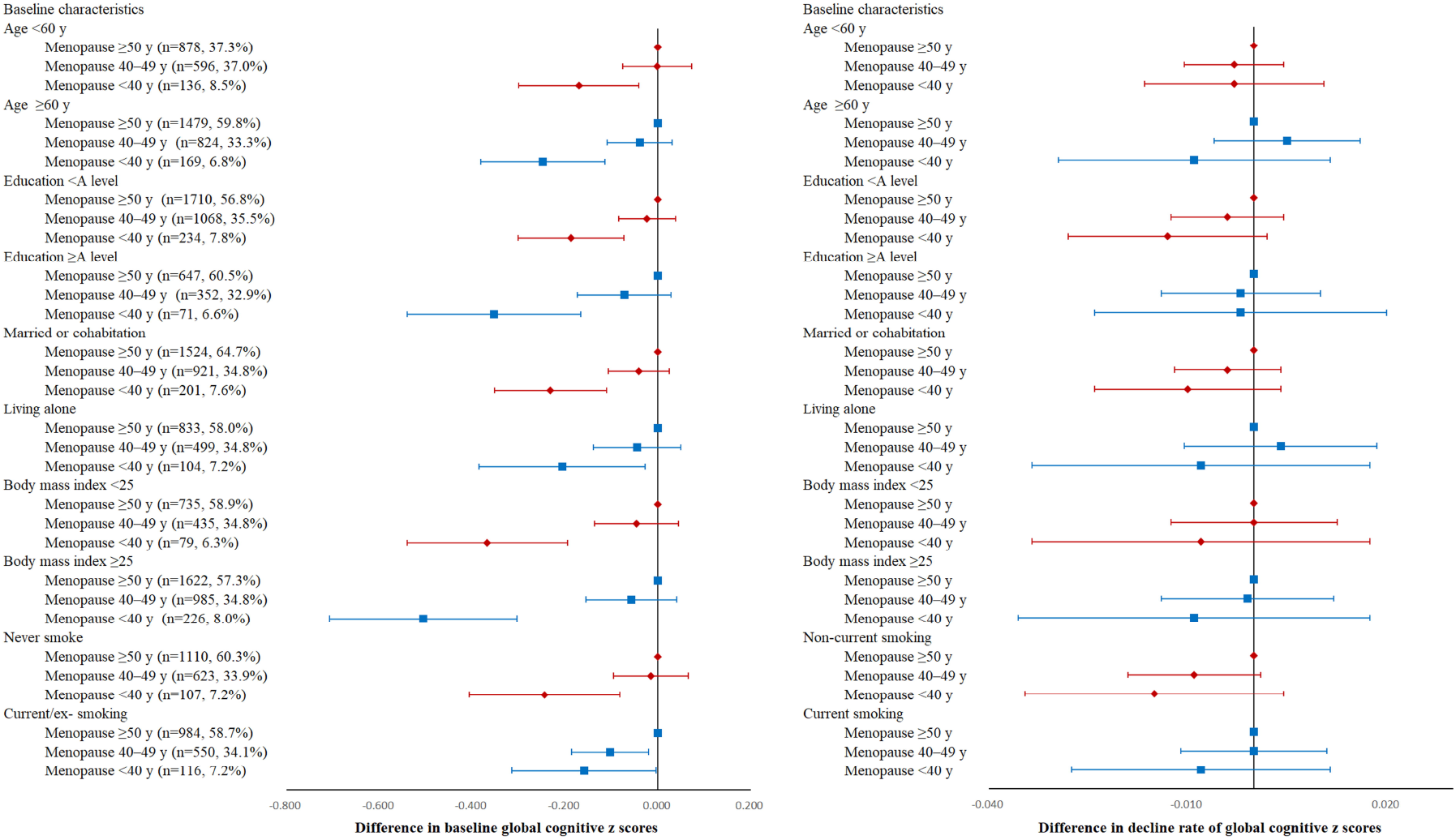
Associations between age at menopause and trajectories of cognitive function in subgroups according to covariates. After adjustment for baseline age, age squared, age×time, BMI, marital status, education level, smoking, drinking, physical activity, depression, hypertension, diabetes, stroke, and cancer, except for the subgroup variable itself.

In sensitivity analysis, including classify age at menopause into tertiles, restricted participants to at least 55 years old at baseline, excluding those who attended study baseline within two years after menopause, and excluding participants with stroke at baseline or during follow-up, the primary results remained consistent. While restricting analysis to participants who received at least 6 cognitive tests, compared with males, females still had higher baseline cognitive scores than males, with a similar long-term rate of decline. In this sensitivity analysis, early menopause was not significantly associated with lower baseline cognitive function (β=-0.076 SD/year, 95% CI -0.191 to 0.040, *p*=0.199; Tables S6–S10).

## 4. Discussion

Using a large prospective cohort with repeated measurements of cognitive function over 14 years, we found that premature menopause (<40 years) was associated with lower cognitive function at baseline compared with menopause >50 years. This association was observed across multiple domains, including global cognition, episodic memory, sematic fluency, and orientation. In contrast, early menopause was not associated with a faster longitudinal decline in global cognition, although it was associated with a faster decline in memory test. Use of HRT or surgery menopause did not modify the association between premature menopause on cognitive trajectories.

Evidence on the association between menopause and long-term cognitive decline remains limited, as most studies focused on risk of dementia or cognitive function at a single time point.[8] In previous studies have repeatedly linked premature menopause ( <4<0 years) to higher risk of dementia or cognitive impairment in both UK and other populations, which is in line with our study.[5, 17, 18] For the other menopause-age, previous results are mixed. A meta-analysis reported a linear association whereby older menopause age was associated with lower dementia risk.[9, 19] A UK Biobank study reported that menopause below the mean (40–49 years) was associated with higher risk, while late menopause (51–54 years) was associated with lower risk.[5] Another UK Biobank study reported an inverted-U shape association in which both menopause <50 years or >50 years increased the risk of dementia.[4] By contrast, a meta-analysis in 2018 reported no association.[20] Among studies assessing cognition at one time point, some reported a linear association,[8] some only reported significant association in premature menopause[21], and one reported that menopause >55 years was associated with lower cognition.[22] We extend prior work by examining the dose–response relationship between age at menopause and cross-sectional/baseline cognitive function. Our RCS analyses suggested an inverted U-shaped association, with menopause around 50 to 55 years associated with the highest baseline global cognitive score, whereas younger or older menopause tended to show lower scores. However, when comparing categories, only menopause <40 years reached statistical significance.

Only a few studies have investigated cognitive decline[22, 23]. An ELSA study reported that compared with menopause >50 years, premature menopause was associated with faster cognitive decline in 2 years. In a China Health and Retirement Longitudinal Study (CHARLS), age at menopause was not associated with cognitive decline during 8-year-follow-up.[22] Our study extend the previous studies by addressing several knowledge gaps. First, the follow-up preriot of previous studies are relatively short. Dementia is a clinical syndrome characterizedby connive decline and it is typically diagnosed once the cognitive performance falls below a defined cut-off point.[24] Evidence showed that cognitive decline may accelerate ten years before dementia diagnosis and drop sharply in a few years before and after diagnosis.[25] Therefore, short follow-up period may have limited power to detect cognitive change related to menopause. Secondly, the UK women, or Caucasian women, had different education level, economic level, and prortion of cognitive impairment. Early studies reported that Chinese women exhibited lower estrogen concentration and shorter reproductive period.[26, 27] In addition, the proportion of premature menopause differs between ELSA and CHARLS.[22] The current study provides a more comprehensive picture of this topic.

Biological mechanisms linking earlier menopause to worse cognitive outcomes remains unelucidated. Some studies attributed these associations to the reduced estrogen exposure after menopause. Estradiol, a principal estrogen, contributes to neurodevelopmental and synaptic processes, has neurotrophic and neuroprotective effects, and may reduce Alzheimer’s disease–related pathology.[3, 11, 28] In addition, younger age at menopause was associated with less gray matter indices (GMV), greater white matter hyperintensities (WMH), and cortical GMV loss, all of which was related to poorer cognitive function.[4]

Considering our primary and subgroup results, we boldly speculate that the adverse impact of premature menopause on cognition may occur primarily in the years shortly after menopause rather than through a persistent, long-term effect, which could contribute to baseline differences observed later. This interpretation is in consistent with our RCS results showing that a longer interval from menopause to baseline was associated with lower baseline scores. Accordingly, future studies should focus on the cognitive health in the years immediately following menopause. Notably, in the subgroup with 19–36 years from menopause to baseline, the observed association with faster decline suggests that the impact of premature menopause may manifest in specific life periods and cognitive domains.[25] Other extensive subgroups were consistent with the primary results, suggesting that the negative effect of premature menopause on baseline cognitive function did not vary substantially across different risk-factor strata.

A number of studies have reported that hysterectomy and bilateral oophorectomy was associated with higher dementia risk.[18, 29, 30] In our analysis, surgical menopause did not modify the effect of age at menopause on cognitive function, suggesting that surgery itself may not be the primary determinant of lower cognitive function. Shorter reproductive duration and reduced lifetime estrogen exposure may be relevant, regardless of whether premature menopause is natural or surgically induced. The effect of HRT on future dementia risk remains controversial. A US cohort of seven thousand females reported that HRT at midlife (40 to 55 years) but not late life reduced dementia risk, supporting the “critical window” hypothesis that timing and duration matter.[31] While the North American Menopause Society guideline that HRT should not be used at any age for preventing dementia.[32] In our study, use of HRT did not modify the effect of premature menopause on lower cognitive function. Hower, we lacke sufficient sample size to evaluate initiation and duration of HRT. Our sensitivity analysis restricting to participants with at least 6 visits represented a healthier subgroup, suggesting that better overall health or greater cognitive reserve could attenuate the negative effect of premature menopause on cognition.

A key strength of this study is that it is the first study evaluating the cognitive trajectories by age at menopause during a long follow-up period. Other strengths include the use of RCS to visualize the potential non-linear association, a large nationally representative UK cohort, and extensive subgroup analyses which may inform preventive strategies.

This study also has limitations. First, age at menopause was self-reported and may be subject to recall bias. However, previous studies have reported that self-reported age at menopause is reasonably reliable, and reliability may be particularly high in UK cohorts.[33] Second, although we hypothesize that accelerated cognitive changes may occur in the years shortly after premature menopause, the mean age in ELSA is relatively older, limiting our ability to capture cognition during the perimenopausal period. Third, we lack some potential confounders, such as APOE genotype and MR measures. Fourth, because the study population was from England, generalizability to other population is a concern. Firth, our study cannot infer causality. Future studies such as Mendelian Randomizationg study are warranted.

In conclusion, we examined cognitive trajectories among UK women according to age at menopause. Compared with menopause >50 years, menopause <40 years was associated with significantly lower cognitive function at baseline. Long-term cognitive trajectories were broadly similar across menopause-age groups. Future studies should focus on cognitive changes in the period shortly after premature menopause, which may contribute to later-life cognitive differences. Surgical menopause and HRT did not appear to exacerbate the negative effect of premature menopause on cognitive function.

## 5. Acknowledgments

The authors thank the U.K. Data Archive for using data from the English Longitudinal Study of Ageing.

## 8. Data availability statement

All data used and analysed in this study are publicly available from the ELSA (https://www.elsa-project.ac.uk). The data that support the findings of this study are available from the corresponding author upon reasonable request.

