## Supplementary Material for "Association between age at menopause and 14-year cognitive trajectories in the English Longitudinal Study of Ageing"

**Contents**

**Table S1. Variables used in this study**

**Table S2. Number of participants in each Waves**

**Table S3. Difference in trajectories of semantic fluency by sex and age at menopause**

**Table S4. Difference in trajectories of memory by sex and age at menopause**

**Table S5. Difference in trajectories of orientation by sex and age at menopause**

**Figure S1. Trajectories of cognitive function in three cognitive tests by sex and age at menopause.**

**Table S6. Sensitivity analysis: difference in trajectories of cognitive function restricted to participants who received at least 6 cognitive assessments**

**Table S7. Sensitivity analysis: difference in trajectories of cognitive function while categorizing age at menopause into tertiles**

**Table S8. Sensitivity analysis: difference in trajectories of cognitive function restricted to participants who were at least 55 years old at baseline**

**Table S9. Sensitivity analysis: difference in trajectories of cognitive function restricted to participants who enrolled in the study at least two years after menopause**

**Table S10. Sensitivity analysis: difference in trajectories of cognitive function after excluding participants who experienced a stroke at baseline or during the follow-up**

**Table S1. Variables used in this study**

| Variable | Data source and question | Codes |
| --- | --- | --- |
| Reproductive factors |  |  |
| Age at menopause | Life history dataset in Wave 3, elsa data in other waves.  Question:  Whether menstruated in last 12 months?  Year had last menstrual period? | rhpee (Wave 3)  hepee (Waves 4, 6-10)  rhpey (Wave 3)  hepey (Waves 4, 6-10) |
| Type of menopause (natural/surgery) | Life history dataset in Wave 3, elsa data in other waves.  Question:  Reason period stopped? | rhper (Wave 3)  heper (Waves 4, 6-10) |
| Age at menarche | Life history dataset in Wave 3, elsa data in other waves.  Question: Age first menstrual period? | rhmen (Wave 3)  hemen (Waves 4, 6-10) |
| Use of hormone replacement therapy (HRT) | Life history dataset in Wave 3, elsa data in other waves. | rhhrt (Wave 3)  hehrt (Waves 4, 6-10) |
| Cognitive function |  |  |
| Memory | Elsa data in Waves 3-5, 7-10. | cflisen  cflisd |
| Orientation | Elsa data in Waves 3-10 | cfdscr |
| Semantic fluency | Elsa data in Waves 3-10 | cfani |
| Baseline characteristics |  |  |
| Age | Elsa data in Wave 3 | dhager |
| Sex | Elsa data in Wave 3 | dhsex |
| Body mass index | Nurse data in Waves 2 and 4 | bmival |
| Marital status | Ifs derived dataset in Wave 3 | marstat |
| Education level | Ifs derived dataset in Wave 3 | qual3 |
| Smoking status | Ifs derived dataset in Wave 3 | smokerstat |
| Alcohol drinking | Elsa data in Wave 3  Question: How often respondent has had an alcoholic drink during the last 12 months? | scako |
| Physical activity | Elsa data in Wave 3  Questions:  Frequency does vigorous sports or activities?  Frequency does moderate sports or activities?  Frequency does mild sports or activities?  Level of physical activity in main job?  0 = inactive; no moderate OR vigorous activity on a weekly basis  1 = mild; (mild activity at least once a week OR moderate activity once a week or less) AND no vigorous activity  2 = moderate; Physical work OR moderate activity at least once a week OR vigorous activity once a week or less  3 = high; heavy manual work OR vigorous activity at least once a week | heacta  heactb  heactc  wpjact |
| Depression | Elsa data in Wave 3  Questions:  Whether felt depressed much of the time during past week?  Whether felt everything they did during past week was an effort?  Whether felt their sleep was restless during past week?  Whether was happy much of the time during past week?  Whether felt lonely much of the time during past week?  Whether enjoyed life much of the time during past week?  Whether felt sad much of the time during past week?  Whether could not get going much of the time during past week?  Cur off point was defined as the 8-item Center for Epidemiologic Studies Depression Scale (CES-D) score$\geq$4. | psceda  pscedb  pscedc  pscedd  pscede  pscedf  pscedg  pscedh |
| Hypertension | Elsa data in Wave 3  Self-reported | hedawbp  hedanbp  hediabp  dhedimbp |
| Diabetes | Elsa data in Wave 3  Self-reported | hedawdi  hedandi  hediadi  dhedimdi |
| Stroke | Elsa data in Wave 3  Self-reported | hedawst  hedanst  hediast  dhedimst |
| Cancer | Elsa data in Wave 3  Self-reported | hedbwca  hedbmca  dhedibca |

**Table 2. Number of participants in each Waves**

|  | Menopause $<$40 y | Menopause 40–49 y | Menopause $\geq$50 y | Men |
| --- | --- | --- | --- | --- |
| Wave 3 | 305 (100%) | 1420 (100%) | 2357 (100%) | 3339 (100%) |
| Wave 4 | 267 (87.5%) | 1290 (90.0%) | 2227 (94.5%) | 3031 (90.8%) |
| Wave 5 | 266 (87.2%) | 1244 (87.6%) | 2109 (89.5%) | 2904 (87.0%) |
| Wave 6 | 246 (80.7%) | 1142 (80.5%) | 1934 (82.1%) | 2653 (79.5%) |
| Wave 7 | 223 (73.1%) | 995 (70.1%) | 1729 (73.4%) | 2318 (69.4%) |
| Wave 8 | 196 (64.3%) | 879 (61.9%) | 1532 (65.0%) | 2035 (61.0%) |
| Wave 9 | 171 (56.1%) | 769 (54.2%) | 1360 (57.7%) | 1774 (53.1%) |
| Wave 10 | 99 (32.5%) | 504 (35.5%) | 891 (37.8%) | 1222 (36.6%) |

**Table S3. Difference in trajectories of semantic fluency by sex and age at menopause^a^**

|  | Difference in baseline  semantic fluency z scores |  | Difference in decline rates of  semantic fluency z scores |  |
| --- | --- | --- | --- | --- |
|  | Coefficient (95% CI) | *p* value | Coefficient (95% CI) | *p* value |
| Sex |  |  |  |  |
| Male | Reference | Reference | Reference | Reference |
| Female | 0.040 (0.001, 0.079) | 0.043 | -0.003 (-0.007, 0.001) | 0.092 |
| Age at menopause |  |  |  |  |
| ≥50 y | Reference | Reference | Reference | Reference |
| 40–49 y | 0.005 (-0.047, 0.058) | 0.840 | -0.007 (-0.013, -0.001) | 0.015 |
| $<$40 y | -0.181 (-0.276, -0.085) | 0.005 | -0.009 (-0.02, 0.001) | 0.076 |

^a^Adjusted for baseline age, age squared, age×time, BMI, marital status, education level, smoking, drinking, physical activity, depression, hypertension, diabetes, stroke, and cancer.

**Table S4. Difference in trajectories of memory by sex and age at menopause^a^**

|  | Difference in baseline  memory z scores |  | Difference in decline rates of  memory z scores |  |
| --- | --- | --- | --- | --- |
|  | Coefficient (95% CI) | *p* value | Coefficient (95% CI) | *p* value |
| Sex |  |  |  |  |
| Male | Reference | Reference | Reference | Reference |
| Female | 0.301 (0.265, 0.337) | <0.001 | -0.002 (-0.005, 0.002) | 0.361 |
| Age at menopause |  |  |  |  |
| ≥50 y | Reference | Reference | Reference | Reference |
| 40–49 y | -0.046 (-0.097, 0.004) | 0.072 | -0.001 (-0.006, 0.005) | 0.751 |
| $<$40 y | -0.133 (-0.227, -0.040) | 0.005 | -0.011 (-0.021, -0.001) | 0.027 |

^a^Adjusted for baseline age, age squared, age×time, BMI, marital status, education level, smoking, drinking, physical activity, depression, hypertension, diabetes, stroke, and cancer.

**Table S5. Difference in trajectories of orientation by sex and age at menopause^a^**

|  | Difference in baseline  orientation z scores |  | Difference in decline rates of  orientation z scores |  |
| --- | --- | --- | --- | --- |
|  | Coefficient (95% CI) | *p* value | Coefficient (95% CI) | *p* value |
| Sex |  |  |  |  |
| Male | Reference | Reference | Reference | Reference |
| Female | 0.107 (0.069, 0.144) | <0.001 | -0.005 (-0.011, 0.001) | 0.104 |
| Age at menopause |  |  |  |  |
| ≥50 y | Reference | Reference | Reference | Reference |
| 40–49 y | -0.011 (-0.062, 0.040) | 0.672 | 0.000 (-0.009, 0.009) | 1.000 |
| $<$40 y | -0.138 (-0.236, -0.039) | 0.006 | 0.002 (-0.015, 0.018) | 0.849 |

^a^Adjusted for baseline age, age squared, age×time, BMI, marital status, education level, smoking, drinking, physical activity, depression, hypertension, diabetes, stroke, and cancer.

**Figure S1. Trajectories of cognitive function in three cognitive tests by sex and age at menopause.** The values were based on the linear mixed models. After adjustment for baseline age, age squared, age×time, BMI, marital status, education level, smoking, drinking, physical activity, depression, hypertension, diabetes, stroke, and cancer. Baseline covariates were set to the most common characteristics: age 65 years, BMI 28, married, education less than o-level, low physical activity, current smoking, alcohol drinking at least once per week, with hypertension, without depression, diabetes, stroke or cancer. Random effects were set to 0. (A) Black line: men; red line: women. (B) Blue line: women with age at menopause $\geq$50 years; orange line: women with age at menopause between 40–49 years. (C) Blue line: women with age at menopause $\geq$50 years; red line: women with age at menopause $<$40 years.

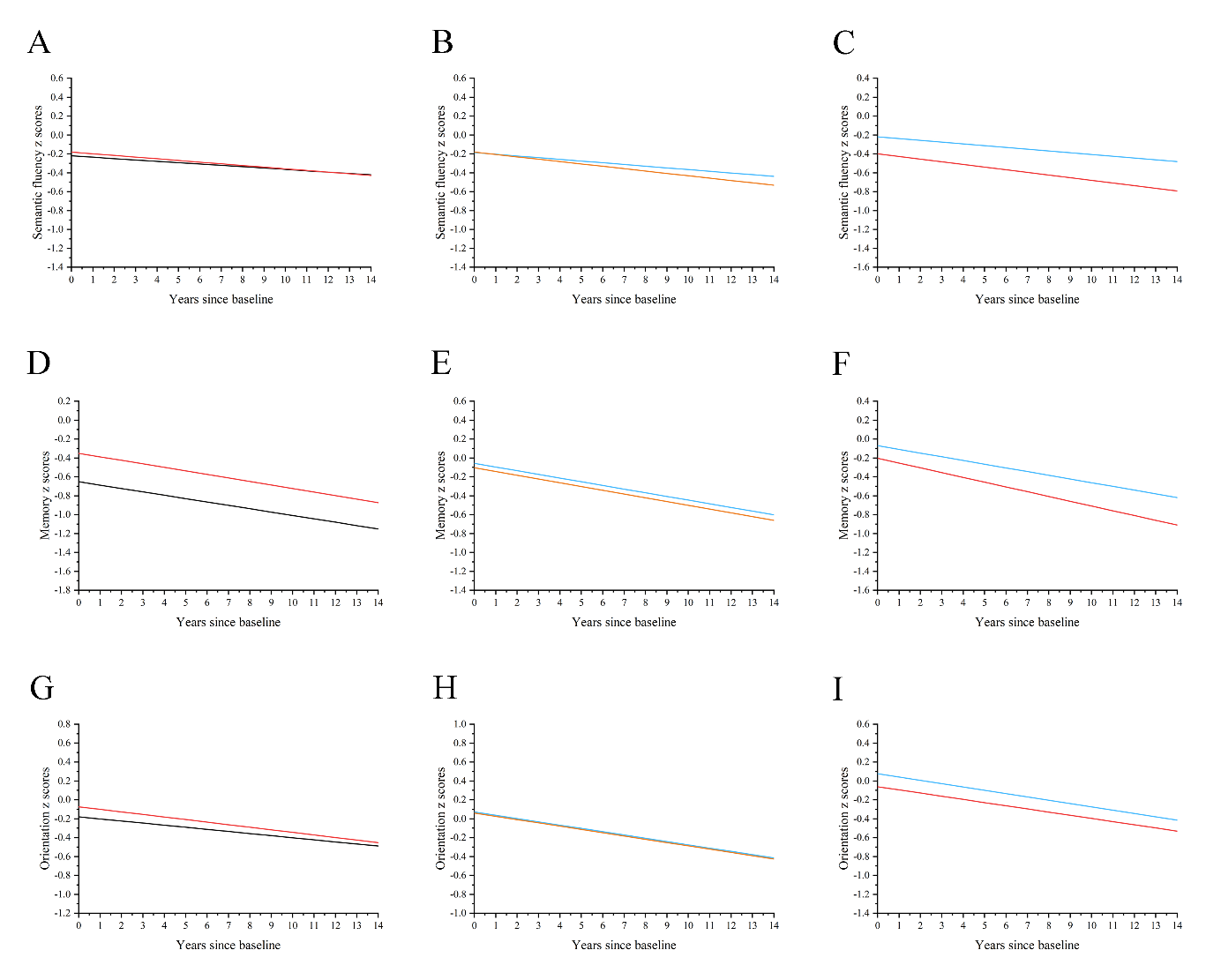

**Table S6. Sensitivity analysis: difference in trajectories of cognitive function restricted to participants who received at least 6 cognitive assessments^a^**

|  | Difference in baseline  global cognitive z scores |  | Difference in decline rates of  global cognitive z scores |  |
| --- | --- | --- | --- | --- |
|  | Coefficient (95% CI) | *p* value | Coefficient (95% CI) | *p* value |
| Sex |  |  |  |  |
| Male (n=1629) | Reference | Reference | Reference | Reference |
| Female (n=2108) | 0.217 (0.172, 0.262) | <0.001 | -0.003 (-0.007, 0.002) | 0.223 |
| Age at menopause |  |  |  |  |
| ≥50 y (n=1262, 59.9%) | Reference | Reference | Reference | Reference |
| 40–49 y (n=703, 33.4%) | -0.032 (-0.094, 0.029) | 0.304 | -0.001 (-0.007, 0.006) | 0.809 |
| $<$40 y (n=143, 6.8%) | -0.076 (-0.191, 0.040) | 0.199 | -0.005 (-0.017, 0.007) | 0.406 |

^a^Adjusted for baseline age, age squared, age×time, BMI, marital status, education level, smoking, drinking, physical activity, depression, hypertension, diabetes, stroke, and cancer.

**Table S7. Sensitivity analysis: difference in trajectories of cognitive function while categorizing age at menopause into tertiles^a^**

|  | Difference in baseline  global cognitive z scores z scores |  | Difference in decline rates of  global cognitive z scores z scores |  |
| --- | --- | --- | --- | --- |
|  | Coefficient (95% CI) | *p* value | Coefficient (95% CI) | *p* value |
| ≥52 y (n=1258, 30.8%) | Reference | Reference | Reference | Reference |
| 48–51 y (n=1335, 32.7%) | -0.037 (-0.094, 0.020) | 0.202 | -0.002 (-0.009, 0.006) | 0.670 |
| $<$48 y (n=1258, 30.9%) | -0.081 (-0.141, -0.020) | 0.009 | -0.003 (-0.011, 0.005) | 0.418 |

^a^Adjusted for baseline age, age squared, age×time, BMI, marital status, education level, smoking, drinking, physical activity, depression, hypertension, diabetes, stroke, and cancer.

**Table S8. Sensitivity analysis: difference in trajectories of cognitive function restricted to participants who were at least 55 years old at baseline^a^**

|  | Difference in baseline  global cognitive z scores |  | Difference in decline rates of  global cognitive z scores |  |
| --- | --- | --- | --- | --- |
|  | Coefficient (95% CI) | *p* value | Coefficient (95% CI) | *p* value |
| Sex |  |  |  |  |
| Male (n=3339) | Reference | Reference | Reference | Reference |
| Female (n=4082) | 0.208 (0.165, 0.250) | <0.001 | -0.005 (-0.011, 0.000) | 0.067 |
| Age at menopause |  |  |  |  |
| ≥50 y (n=2357, 57.7%) | Reference | Reference | Reference | Reference |
| 40–49 y (n=1296, 31.8%) | -0.047 (-0.106, 0.011) | 0.112 | 0.004 (-0.005, 0.012) | 0.414 |
| $<$40 y (n=429, 10.5%) | -0.239 (-0.348, -0.130) | <0.001 | -0.004 (-0.019, 0.011) | 0.638 |

^a^Adjusted for baseline age, age squared, age×time, BMI, marital status, education level, smoking, drinking, physical activity, depression, hypertension, diabetes, stroke, and cancer.

**Table S9. Sensitivity analysis: difference in trajectories of cognitive function restricted to participants who enrolled in the study at least two years after menopause^a^**

|  | Difference in baseline  global cognitive z scores |  | Difference in decline rates of  global cognitive z scores |  |
| --- | --- | --- | --- | --- |
|  | Coefficient (95% CI) | *p* value | Coefficient (95% CI) | *p* value |
| Age at menopause |  |  |  |  |
| ≥50 y (n=1885, 54.6%) | Reference | Reference | Reference | Reference |
| 40–49 y (n=1132, 33.3%) | -0.052 (-0.109, 0.005) | 0.077 | -0.001 (-0.009, 0.007) | 0.807 |
| $<$40 y (n=409, 12.0%) | -0.256 (-0.360, -0.153) | <0.001 | -0.012 (-0.026, 0.002) | 0.082 |

^a^Adjusted for baseline age, age squared, age×time, BMI, marital status, education level, smoking, drinking, physical activity, depression, hypertension, diabetes, stroke, and cancer.

**Table S10. Sensitivity analysis: difference in trajectories of cognitive function after excluding participants who experienced a stroke at baseline or during the follow-up^a^**

|  | Difference in baseline  global cognitive z scores |  | Difference in decline rates of  global cognitive z scores |  |
| --- | --- | --- | --- | --- |
|  | Coefficient (95% CI) | *p* value | Coefficient (95% CI) | *p* value |
| Age at menopause |  |  |  |  |
| ≥50 y (n=1678, 54.9%) | Reference | Reference | Reference | Reference |
| 40–49 y (n=1024, 33.5%) | -0.039 (-0.091, 0.012) | 0.135 | -0.004 (-0.011, 0.003) | 0.222 |
| $<$40 y (n=357, 11.7%) | -0.207 (-0.305, -0.109) | <0.001 | -0.004 (-0.017, 0.008) | 0.503 |

^a^Adjusted for baseline age, age squared, age×time, BMI, marital status, education level, smoking, drinking, physical activity, depression, hypertension, diabetes, and cancer.
